# Wearing masks and establishing COVID-19 areas reduces secondary attack risk in nursing homes

**DOI:** 10.1101/2020.11.27.20239913

**Authors:** Bastien Reyné, Christian Selinger, Mircea T Sofonea, Stéphanie Miot, Amandine Pisoni, Edouard Tuaillon, Jean Bousquet, Hubert Blain, Samuel Alizon

## Abstract

**Background:** COVID-19 is spreading rapidly in nursing homes (NHs). It is urgent to evaluate the effect of infection prevention and control (IPC) measures to reduce COVID spreading.

**Methods:** We analysed COVID-19 outbreaks in 12 NH using rRT-PCR for SARS-CoV-2. We estimated secondary attack risks (SARs) and identified cofactors associated with the proportion of infected residents.

**Results:** The SAR was below 5%, suggesting a high efficiency of IPC measures. Mask-wearing or establishment of COVID-19 zones for infected residents were associated with lower SAR.

**Conclusions:** Wearing masks and isolating potentially infected residents appear to limit SARS-CoV-2 spread in nursing homes.

## Introduction

COVID-19 is spreading rapidly to nursing homes (NH) [1-3] and recommendations have been issued to reduce new cases in NH where a COVID-19 outbreak has been identified [4]. Guidelines from the European Geriatric Medicine Society (EuGMS) mention a variety of interventions based on testing, mask-wearing, or isolation of people with established or suspected infection and their contacts [5]. The relative impact of the different interventions on the magnitude of potential COVID-19 outbreaks in NH remains unclear and requires testing.

In France, the COVID-19 epidemic wave is estimated to have started mid-January 2020 [6]. The first COVID-19 cases in a French NH were detected in the Hérault department (France) on March 10. This first outbreak triggered an immediate response from the Regional Health Authority (ARS Occitanie). Infection prevention and control (IRC) guidelines were implemented and included mask-wearing, establishment of “COVID-19 units” to isolate exposed or infected residents (Table S1 online), and repeated testing for SARS-CoV-2 [2]. Several studies have assessed the secondary attack risk (SAR) of SARS-CoV-2, showing, for instance, that mask-wearing reduces virus transmission in households [7-9]. However, we are not aware of similar studies in NH.

The goal of this study was to investigate the efficiency of IRC measures implemented in the Hérault department (Occitanie region, France) in reducing the spread of SARS-CoV-2 in NHs when a patient was tested positive. We first estimated the SAR defined as the proportion of individuals infected (positive rRT-PCR) in a NH after an outbreak [10, 11]. We further analysed the data using classical statistical modelling to better understand the relative role of specific factors.

## Methods

### Survey

In the Hérault department (France), an observational retrospective longitudinal study was carried out in 12 NH which experienced a COVID-19 outbreak between March and May 2020.

After clinical identification of a COVID-19 case in any NH, all residents were tested via rRT-PCR on nasopharyngeal swab tests. COVID-19 IPC measures were applied to all residents who were clinically followed up for 6 weeks, with repeated PCR testing. Serum antibodies were assessed at the end of the survey by two clinical laboratories that performed the analyses. Blood testing for IgG antibodies directed against the SARS-CoV-2 nucleocapsid protein used an enzyme-linked immunosorbent assay CE-IVD marked kit (ID screen SARS-CoV-2-N IgG indirect from IDVet, Montpellier, France). In the following analyses, positivity was based on rRT-PCR tests. As reported earlier, there was a 95% match between RT-PCR and the serological result [2].

From March 3, all the staff members and residents of all NH agreed to participate in the study. This observational study was approved by the Internal Review Board from the Montpellier University Hospital (IRB-MTP_2020_06_202000534).

Seven main factors associated with each NH were extracted from a survey carried out among the directors of the 12 NH:

- number of floors,
- number of days between the first COVID-19 case and the generalisation of mask-wearing,
- number of medical staff per resident,
- presence or not of a “COVID unit”, *i*.*e*. isolation of infected patients,
- reported sufficient or lack of mask availability after the detection of the first case,
- proportion of single rooms,
- presence or not of temporary agency workers before the detection of the first case.

#### Statistical analyses

We first estimated the Secondary Attack Risk (SAR) of the selected outcomes using a model-based approach described by Bailey [10]. Formally, the probability that *j* persons have been infected among *n* susceptible persons knowing there were *a* introduction events in the household during the outbreak is given by the formula

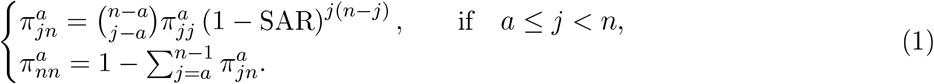

Here, the “household” is assumed to be the floor of a NH. The value of the SAR was estimated using a maximum likelihood approach.

By treating the proportion of infected residents in each NH after the outbreak (*f*) as a final epidemic size, we estimated the basic reproduction number (denoted ℛ_0_) in these NHs by solving the following classical equation [12]:

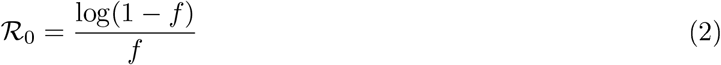

To study the effect of the 7 main variables on the proportion of infected residents, we used generalized linear models (GLMs) with a binomial distribution for the response variable weighted by the total number of residents. In these models, the unit of analysis (the “household” in the SAR model) is the NH. We performed GLMs with all possible combinations of our 7 factors (*i*.*e*. 2 ^7^ = 128 models) and performed model selection using the Akaike Information Criterion (AIC). An AIC difference of 2 between models was considered to be significant following classical statistical practice [13].

To improve statistical power, we also performed the analysis at the level of NH floors. The rationale for accounting for such structure within NHs is that activities between groups of residents were cancelled and that French national guidelines incited NHs to separate different structures and staff. To partly correct for non-independence issues, we used a nested structure with the floors being associated to a NH, which was itself treated as a random effect in the model. This was done using Generalised Linear Mixed Models (GLMMs) with a binomial distribution for the response variable and a normal distribution for the random effect [14]. As for GLMs, the model comparison was performed using AICs. Analyses were done using the lme4 package (`glmer` function) [15] in `R` version 4.0.2.

## Results

The study involved 930 residents and 360 medical staff spread over 40 floors from 12 nursing homes (NHs), with 3.3 floors on average per NH. Details regarding the nursing homes can be found in Table SI. The first rRT-PCR positive cases were detected on March 10, 2020, while the last NH of the area to be affected by the first wave reported its first case on April 21, 2020.

Assuming a single virus introduction per NH floor and independence between the floors, we estimated a SAR of 0.041 (95% CI: [0.036,0.047]) among the residents. Assuming two virus introductions instead of one per NH floor decreased the estimate to 0.033 (95% CI: [0.028,0.038]). Based on the final outbreak size, i.e. the total number of residents infected per NH floor, we found that the estimate for the basic reproduction number (ℛ_0_), assuming a single introduction per NH, was 1.021 (95% CI: [1.018,1.024]).

We then used GLMMs to identify the factors associated with NH outbreak size. The GLMM with the lowest Akaike Information Criterion (AIC), shown in Table 1, contained two factors, the delay in maskwearing (in days) and the reported mask-availability. These factors were both significantly associated with larger outbreaks as shown in Figure S1. When analysing the 11 GLMMs that were comparable from a statistical point of view (their differences in AIC with the best model was smaller than 2), we found consistent results. Table 2 shows that the two co-variables identified in the best model were the most often significant in the 11 models. Another significant co-factor was the presence of a “COVID unit”, which decreases outbreak size. Finally, in some of the models, the presence of temporary agency workers before the first case was associated with larger outbreaks.

**Table 1:** Effect of significant factors on outbreak size in NH. This generalised linear mixed model was selected using an AIC criterion.

| Fixed effect | Estimate | Std. Err. | z-value | Pr(> \|z\|) |
| --- | --- | --- | --- | --- |
| (Intercept) | -2.38 | 0.40 | -6.03 | 1.6e-09 |
| delay in mask wearing | 0.59 | 0.19 | 3.08 | 0.0021 |
| enough masks | 1.35 | 0.44 | 3.05 | 0.0023 |

**Table 2:** Co-factors identified in the 11 best GLMM. We show the proportion of models that contain this co-factor, the proportion of models in which it is significant (p-value<0.05 in the GLMM), and its effect on the total number of infection if significant.

| Co-factor | Proportion | Prop. Signif. | Effect |
| --- | --- | --- | --- |
| Days until mask wearing | 0.91 | 0.73 | + |
| Report enough masks | 0.82 | 0.73 | + |
| COVID-unit | 0.55 | 0.18 | − |
| Part-time workers | 0.36 | 0.18 | + |
| Medical staff | 0.18 | 0 | NA |
| Prop. single room | 0.18 | 0 | NA |
| Floors | 0.09 | 0 | NA |

When assuming a less detailed model without any structure at the floor level, we identified 11 generalised GLMs that performed comparatively well from a statistical point of view (their AIC difference with the best model was smaller than 2). The presence of a COVID-unit had a significant effect in all 11 models. The next significant effects were the presence of temporary agency workers before the first case (10 models out of 11), the delay in mask-wearing (6 models out of 11) and, for two models, the number of floors in the NH. The effect of the factors on outbreak size was the same as for the GLMMs.

## Discussion

First, we estimated the SAR of COVID-19 outbreaks in NH and found values lower than 5%., which is much lower than earlier estimates that ranged from 13.8%. to 19.3%. [8], 23%. [9], or even 35%. [7]. This is consistent with the strict IPC measures implemented after the first outbreak was detected on March 10, 2020.

Second, we conducted statistical modelling analysis to identify the relevant cofactors that best explain the heterogeneity of NH spreading of COVID-19. Generalised Linear Models (GLM) were used because the response variable was not continuous (number of infected residents per NH), making classic linear regression such as ANOVA or ANCOVA inadequate. The structure of the data allowed us to gain even more insight by working at the floor level. However, this raises non-independence issues between the same floor of a NH. To address this, we use Generalised Linear Mixed Models (GLMM), also known as hierarchical generalized linear models, which are commonly used in clinical research [14]. In these model, the NH as a ‘random’ effect thereby partly corrected for the non-indcpcndcncc issue, although care had to be taken in model interpretation [15].

Our main approach was to analyse epidemics at the level of a NH floor using GLMMs. This was motivated by the fact that early guidelines led to contact limitations in NH (e.g. cancellation of group events, or assignment of members of the staff to specific floors). We found that two factors affected the epidemic spread within the NHs: the delay in mask-wearing and the reported mask availability. The earlier mask-wearing was generalised in the NH, the smaller the outbreak. Unexpectedly, reporting a lower mask availability was associated with fewer outbreaks. While this effect should be handled with care because of the subjective dimension of the variable, an explanation could be that a (reported) shortage of masks occurred in the NHs that were using more masks.

Interestingly, when using less detailed statistical models that ignored the floor structure (i.e. GLMs), therefore assuming that cases occurred homogeneously in the NH, the main effect we found was the setting up of a “COVID unit”, which was associated with smaller outbreaks. This further strengthens our choice to use a detailed GLMM model to analyse the data. The presence of temporary agency workers before the first case was also associated with larger outbreaks in some of the GLMs.

One potential limitation of the analysis could be the presence of over-dispersion in the data. When correcting for this potential bias, only 9 of the 127 potential GLMMs had a significant factor, which was the presence of a COVID-unit in the NH. Unfortunately, correcting for overdispersion requires the use of quasi-likelihoods, therefore precluding AIC-based model comparison.

Another major limitation of this study is that it was conducted retrospectively, which constrained the design and variable choices. In particular, some of the variables are subjective, which can generate unexpected correlations. However, this work can be considered as a pilot and will help to design further prospective studies with improved statistical power and the possibility to potentially include additional factors in the analysis.

Overall, these results confirm the efficiency of the measures implemented in the South of France to prevent SARS-CoV-2 epidemics in nursing homes. They reveal the importance of within-NH structure. They also show that delays in the generalisation of mask-wearing before the first case are strongly associated with the magnitude of the outbreaks. Finally, they support the US CDC and European guidelines, which both recommend that NH facing a COVID-19 outbreak dedicate an area of the facility with specific staffing and ICP measures to the care of residents with suspected or confirmed COVID-19 infections [2, 4].

## Data Availability

Raw data will be published along with the article when accepted.

## Footnotes

### Presented in part

Covid-19 Dynamics & Evolution, October 19-20, 2020 (online meeting).

## Acknowledgements

The authors would like to thank the residents and their families for participating in the study, along with all of the NH staff for taking the time to answer a survey, while helping their residents during the crisis. The authors also thank IdVet for providing serological tests and Anna Bedbrook for her help in improving the writing.

---

**Figure SI:**
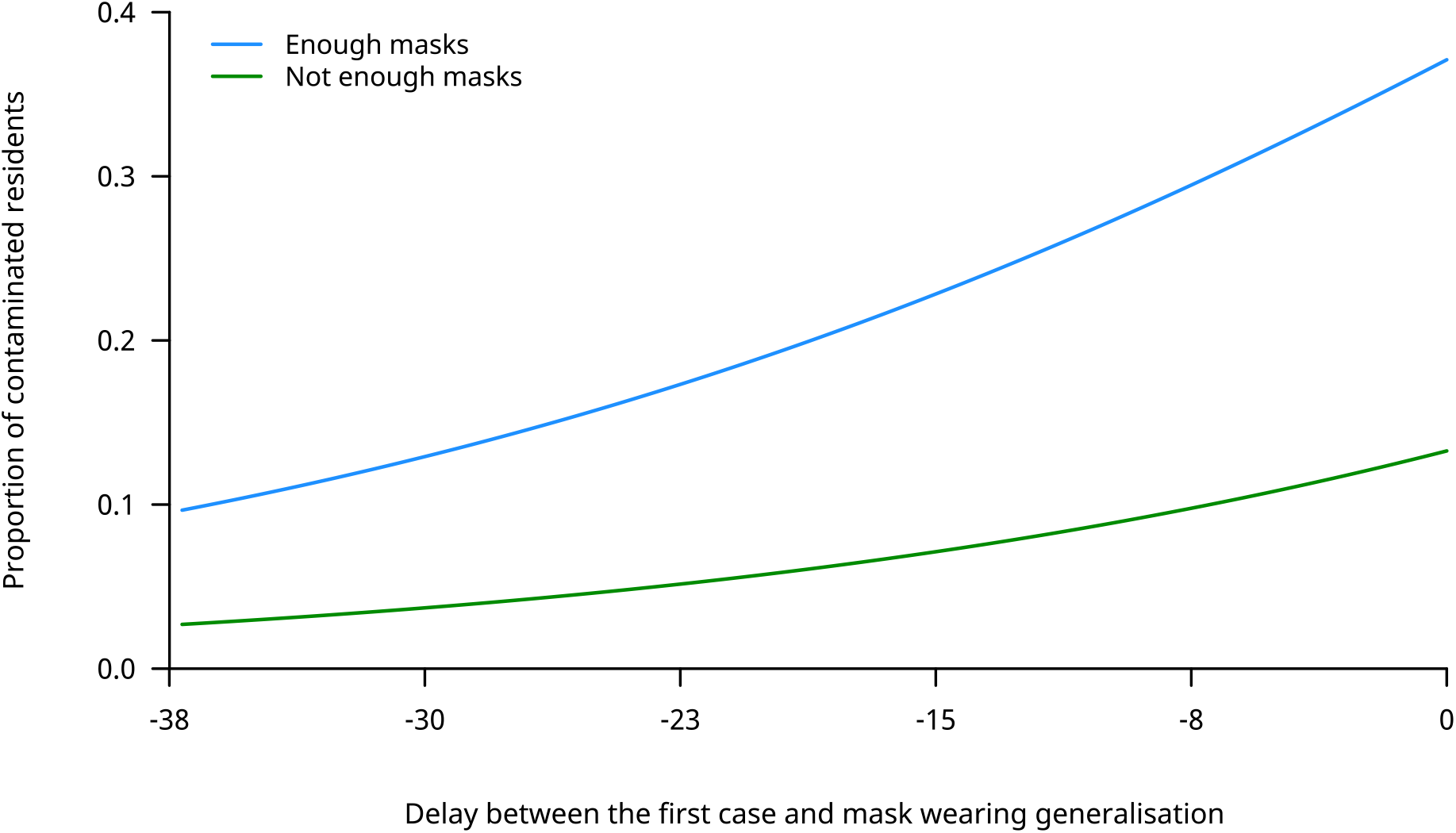
Effect of delay in mask-wearing and mask availability on the proportion of infected residents. The plot is based on the coefficients from the GLMM model shown in Table 1.

**Table SI:**
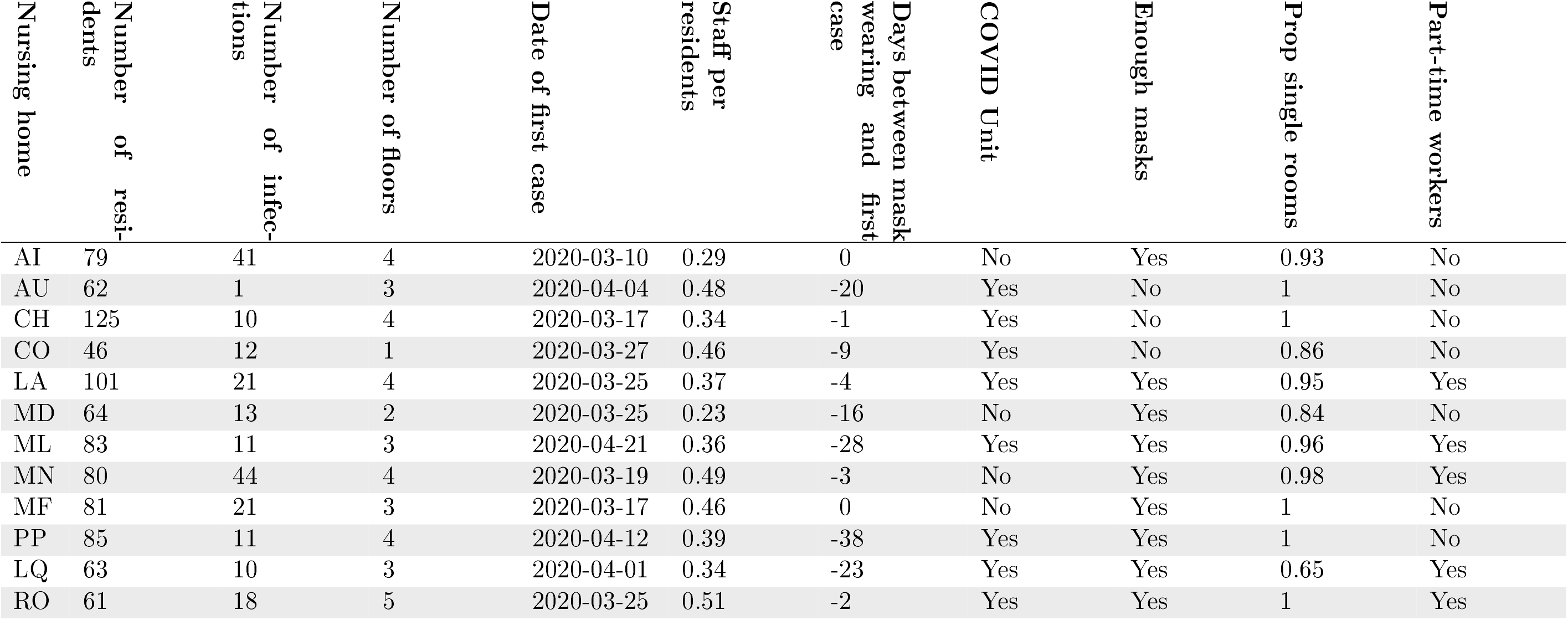
Nursing home details. The proportion of infected staff and residents were estimated using serological assays. Missing data are indicated with ‘NA’.

